# DEVELOPMENT AND VALIDATION OF AN ENZYME-LINKED IMMUNOASSAY KIT FOR DIAGNOSIS AND SURVEILLANCE OF COVID-19

**DOI:** 10.1101/2021.06.23.21259392

**Authors:** Flávia F. Bagno, Sarah A. R. Sérgio, Maria Marta Figueiredo, Lara C. Godoi, Luis A. F. Andrade, Natália C. Salazar, Camila P. Soares, Andressa Aguiar, Flávia Jaqueline Almeida, Edimilson D. da Silva, Antônio G. P. Ferreira, Edison Luiz Durigon, Ricardo T. Gazzinelli, Santuza M. R. Teixeira, Ana Paula S. M. Fernandes, Flavio G. da Fonseca

**Author notes:** Correspondence: Correspondence: / / 55-31- 34092746. These authors contributed equally to this work (Ana Paula S. M. Fernandes, Flavio G. da Fonseca).

## Abstract

There is a massive demand to identify alternative methods to detect new cases of COVID-19 as well as to investigate the epidemiology of the disease. In many countries, importation of commercial kits poses a significant impact on their testing capacity and increases the costs for the public health system. We have developed an ELISA to detect IgG antibodies against SARS-CoV-2 using a recombinant viral nucleocapsid (rN) protein expressed in *E. coli*. Using a total of 894 clinical samples we showed that the rN-ELISA was able to detect IgG antibodies against SARS-CoV-2 with high sensitivity (97.5%) and specificity (96.3%) when compared to a commercial antibody test. After three external validation studies, we showed that the test accuracy was higher than 90%. The rN-ELISA IgG kit constitutes a convenient and specific method for the large-scale determination of SARS-Cov-2 antibodies in human sera with high reliability.

## 1. Background

SARS-CoV-2, a member of the *Coronaviridae* family, *Betacoronavirus* genus, is the causative agent of COVID-19, a disease marked by the occurrence of severe acute respiratory syndrome in many of the infected patients. The virus was first isolated on December, 2019, in Wuhan, China. Since then, it has spread quickly, and, by March 11, 2020, the World Health Organization (WHO) declared COVID-19 pandemic. As of June 2021, the disease is close to reach the astonishing mark of 170 million people affected worldwide and has caused more than 3.7 million deaths [1]. Emergency of virus variants has brought more complexity to this scenario, since they may have been associated to overwhelmingly new outbreaks in large cities such as Manaus, Brazil [2].

Although the combination of PCR and serological tests such as enzyme-linked immunosorbent assays (ELISA) are ideal for an accurate diagnosis, the detection of antibodies is particularly relevant during later stages of infection [3]. Taking into account the rate of asymptomatic infections and the extent of the disease transmission, serologic assays are needed for epidemiologic studies and surveillance. Also, serological tests may identify individuals who have developed immunity after the disease and are highly relevant as an assessment tool after vaccination [4–6].

SARS-CoV-2 has four structural proteins: Spike (S,) Envelope (E), Membrane (M), and Nucleocapsid (N). The S protein, which is cleaved into S1 (containing the receptor binding domain, RBD) and S2 (further cleaved into S2’ to form the viral fusion peptide) subunits is critical for viral entry and has been described as a neutralizing target [7,8]. Besides the S protein, the SARS-CoV-2 N protein has been described as an immunodominant antigen in studies with COVID-19 patients. Similar to SARS-CoV-1 (the causative agent of the 2003 SARS) N protein, this antigen is highly expressed during infection [9–11], making it a suitable antigen to be included in serology-based diagnostics [13-15].

Due to the urgency and high demand, many serological tests to diagnose COVID-19 have been rapidly developed-and made available on the market, with validation often limited to testing relatively few clinical samples [15], and some commercial ELISA kits have been validated worldwide [16] and approved by health regulatory agencies. However, none of the currently available serological kits are fully nationalized in several countries, which makes the products more expensive, less accessible, and subject to availability fluctuation due to international demands and importation difficulties. This is particularly critical in Brazil, considering that the country is the third in the world in number of cases, and the second in terms of death toll [1].

We aimed to developed a new ELISA kit to help COVID-19 diagnosis (henceforth named EIE COVID-19 IgG kit) through the detection of IgG antibodies against the SARS-CoV-2 nucleocapsid (N) protein.

## 2. Study design

### 2.1. Antigen production

The full length coding region of the nucleocapsid (N) gene of SARS-CoV-2 (Genebank accession number: MT126808.1) was codon-optimized, subcloned into pET-24a-(+) expression vector and used to transform *E.coli* BL21(DE3) strain. Plasmid-positive clones were cultivated in LB medium, cultures were induced with IPTG (0,5 mM, 4h) and the recombinant protein expression levels were examined by SDS-PAGE. The recombinant antigen was purified by affinity chromatography using nickel columns in an AKTAprime plus system following manufacturer’s instructions (GE Healthcare, USA).

### 2.2. Sera bank and ethical considerations

The use of sera samples from patients and healthy volunteers was approved by the UFMG’s Ethics Committee and by the National Research Ethics’ Committee (CAAE: 1686320.0.0000.5149). Negative sera obtained before 2020 are from healthy donors, and sera obtained after 2020 are from individuals who tested negative for the viral RNA screening test qRT-PCR (nasal swab). Positive samples were selected based on a reported history of a positive SARS-CoV-2 PCR nasal swab, or on results from a rapid test-dual path platform (DPP) COVID-19 IgM/IgG, according to the supplier’s instructions (Bio-Manguinhos, Fiocruz, Brazil). Considering samples from patients confirmed by positive qPCR result (+qPCR) and used during the internal validation (n=54), 20,4% (n=11) were from hospitalized individuals and 79,6% (n=43) were from mildly symptomatic, non-hospitalized patients. No clinical information about the hospitalized patients were provided. Information on positive mildly symptomatic patients is available (Supplementary data Table S1). The median time between the PCR results and the first blood collection (out of serial collections) was 7 (± 1) days.

In all, 894 samples were tested, 362 from SARS-CoV-2-positive patients (240 individuals), 407 from SARS-CoV-2-negative donors and 125 samples from patients bearing other different conditions (patients that were seropositive for other viruses, homolyzed sera, icteric sera and sera with increased levels of rheumatoid factors). Figure 1 summarizes how samples were used for the rN-ELISA kit validation.

**Figure 1.**
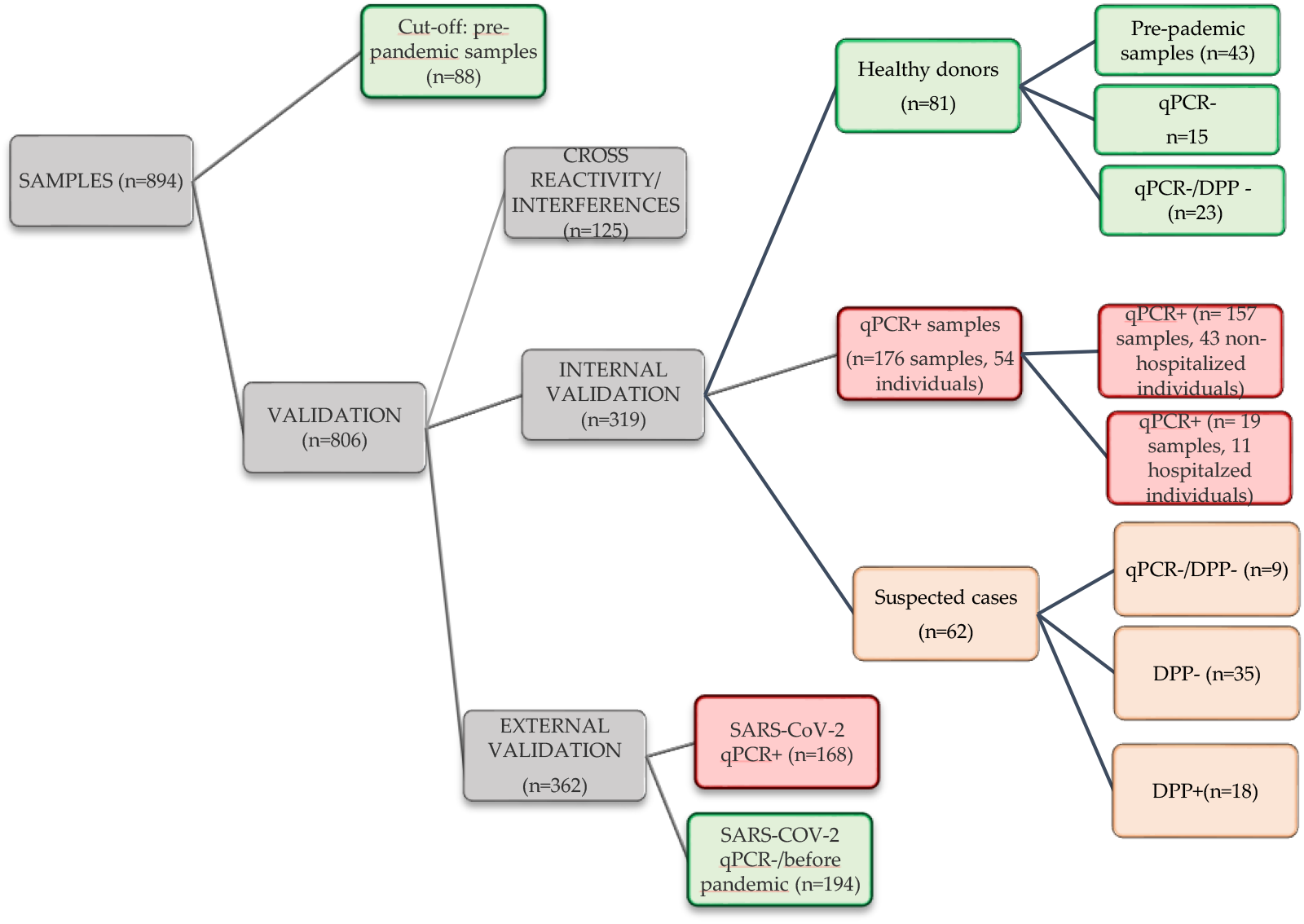
Sera samples used during rN-ELISA validation. Experiments are categorized and showed in gray, positive samples (qPCR or DPP) in red, negative samples (negative PCR or pre-pandemic) in green and suspected cases (negative or no PCR data, but that had contact with confirmed COVID-19 individuals) in orange. The rN-ELISA sensitivity and specificity during internal validation included samples from healthy donors (n=81) and qPCR positive samples from hospitalized (n=11) and non-hospitalized individual (n=43). Agreement between rN-ELISA and DPP included: healthy donors tested by DPP (n=23), all qPCR+ samples (n=176) and suspected cases(n=62). The positive agreement to qPCR was calculated using samples from 43 non-hospitalized individuals (n= 157 samples, with information about the time after the qPCR confirmation).

### 2.3. ELISA

The ELISA was performed by coating plates of polystyrene (Costar, USA) with the recombinant antigen diluted in carbonate buffer to a final concentration of 4 μg/mL (overnight at 4 °C). Wells were blocked (PBS with 1% bovine serum albumin) for 2h at room temperature (25 ± 2°C). For each assay, samples diluted (1:101) in PBS-T (0.05% Tween-20) were added and incubated for 30 min at 37°C. After five washes (PBS-T, 1% Tween-20), the conjugate (1:5.000 in diluent Moss, horseradish peroxidase HRP-conjugated anti-human IgG goat immunoglobulin, Fapon, China) was added and the plates were further incubated (30 min at 37°C). After further washing, reactions were revealed using TMB (3,3’,5,5;-tetramethylbenzidine, Moss, USA) for 15 min, and H_2_SO_4_ (0,5 M) was added to stop reactions. Plates were analyzed in a Microplate Reader at optical density (OD) of 450 nm.

A cut-off value was determined using 88 samples from healthy donors (obtained before 2020) according to equation 1:

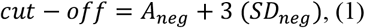

*A*_*neg*_: Average of negative samples (OD)

*SD*_*neg*_: standard deviation of negative samples (OD)

Positive and negative controls were prepared using heat-inactivated samples (56°C, 30 minutes) [17].

The cut-off of the following tests was based on the positive control (equation 2):

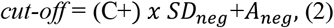

The internal evaluation of accuracy included the testing of 135 serum samples, from patients admitted to a hospital in Belo Horizonte (Minas Gerais, Brazil) and presenting positive PCR for COVID-19 (n=11); sera from non-hospitalized individuals (symptomatic or oligosymptomatic) presenting positive PCR for COVID-19 (n=43, 13±2,2 days post PCR confirmation); sera from healthy donors (taken after 2020) who tested negative for SARS-CoV-2 nasal swab PCR (n= 38); and sera from healthy donors who sampled blood before the COVID19 emergence (before 2020) (n=43). P values were determined through unpaired, two-sided Mann–Whitney U-test. To all validation assays, an index (*I*) for each sample was calculated, according to equation 3:

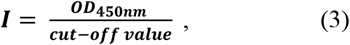

The results were classified as: non-reactive (I<0.8), borderline (0.8≤ *I* <1.1) or reactive (I≥1.1).

## 3. Results

### 3.1. rN-ELISA accuracy

The full-length recombinant N protein (MW 48 KDa), was expressed at high yields in *E. coli* BL21(DE3) (60 mg of purified protein/mL of bacterial culture), and obtained at high purity grade (data not shown). The cut-off value for the optical density obtained with the rN-ELISA test was defined as three standard deviations greater than the average OD450 of 88 negative samples (obtained before 2020, from healthy individuals) (cut-off=0.426, Supplementary materials, Table S2). Besides these samples, 81 negative sera were tested (before 2020, or after 2020 with PCR negative results with two-weeks intervals) (Supplementary materials, Table S3), 76 of them presented a non-reactive result (Index<0.8, with Index, or *I*=OD_450nm_/cut-off), two presented a reactive result (*I*>1.1) and three were classified as borderline samples (0.8≤ *I* <1.1), corresponding to a 96.3% (95% CI: 89.7-99.0%) specificity for the test. Of all samples that tested positive by qRT-PCR (n=54, 13±2 days post molecular confirmation), 44 presented positive results (*I* ≥ 1.1), four were classified as borderline samples, and six presented negative results (Supplementary materials, Table S4). These resulted on a sensitivity of 83,3% (CI: 71.3-91.0%) for the assay. Sensitivity increased to 97.5% (87.1-99.9) % when we considered rapid test DPP-confirmed positive samples (38/40, two borderline samples). Results were further supported by ROC curves (Figure 2).

**Figure 2.**
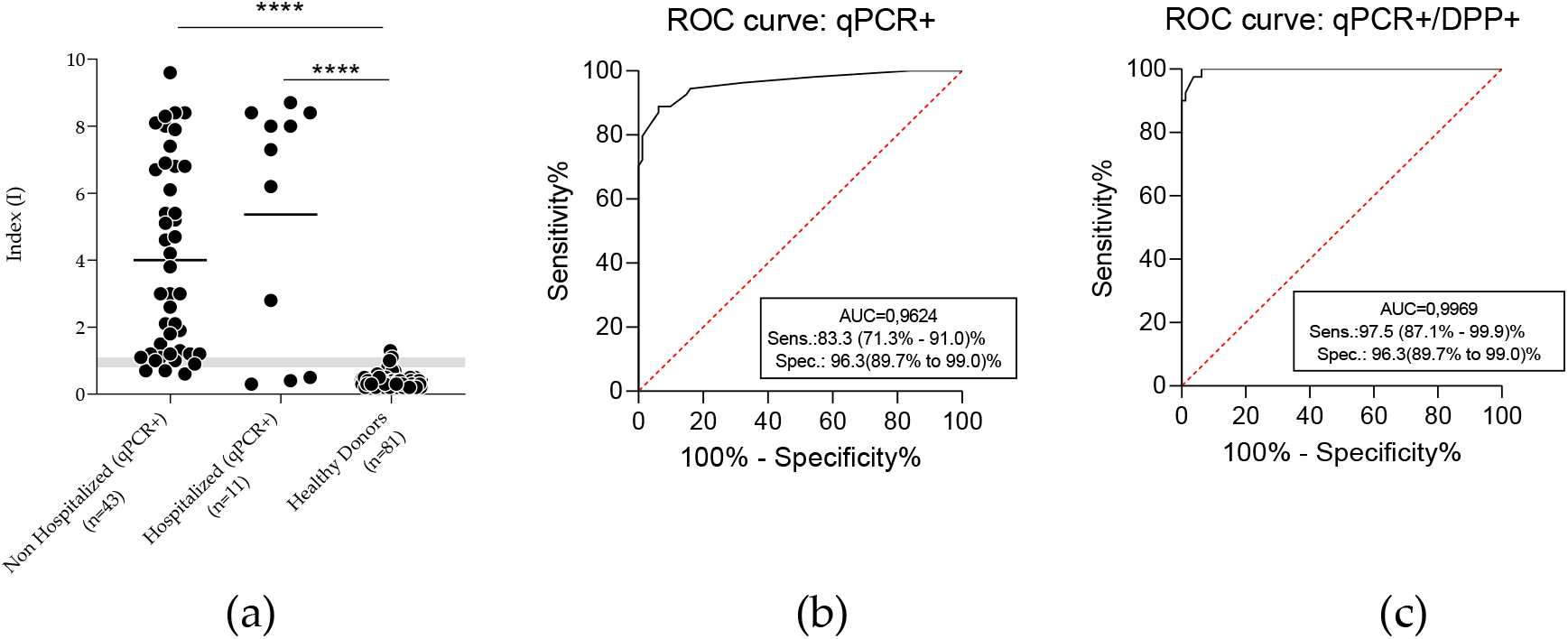
Evaluation of the accuracy of anti-SARS-CoV-2 rN-ELISA IgG kit. (a) The sensitivity and specificity of the rN antigen were calculated according to the index in ELISA and confirmed by ROC curve. A comparison between qPCR positive patients and healthy donors showed significant differences between groups (p<0,0001, by Mann Whitney test). Grey zone (Index ranging from 0.8-1,09) indicates borderline results. (b) ROC curve considering all PCR positive results, including antibody non detected by DPP. (c) Analysis considering PCR positive patients with positive antibody confirmation by DPP (serological reference test).

### 3.2. Cross-reactivity (analytical specificity) and interferences

A cross-reactivity study was performed with a panel of clinical samples from patients infected with other, non-SARS-CoV2, relevant human viruses. Low cross-reactivity (1/19) was observed with influenza antibody-positive samples. Cross-reactions to other human pathogenic viruses were not observed. Negative samples with high levels of rheumatoid factors, hemoglobin and bilirubin were also tested and did not cause any detectable interference in the rN-ELISA IgG kit detection capability (Table 2).

**Table 2.** Sera bank tested with rN-ELISA IgG kit in cross-reactivity and interference studies.

| PANEL | N | rN-ELISA IgG kit |  |
| --- | --- | --- | --- |
|  |  | Negatives | % Negatives |
| Fresh vaccination against Influenza<sup>1</sup> | 8 | 8 | 100% |
| Influenza antibody positive<sup>2</sup> | 19 | 18 | 95% |
| Measles antibody positive<sup>2</sup> | 15 | 15 | 100% |
| Measles PCR positive<sup>3</sup> | 10 | 10 | 100% |
| Human Parvovirus antibody positive<sup>3</sup> | 6 | 6 | 100% |
| Chikungunya antibody positive<sup>1</sup> | 8 | 8 | 100% |
| Dengue antibody positive<sup>1</sup> | 8 | 8 | 100% |
| Zika antibody positive<sup>1</sup> | 8 | 8 | 100% |
| Yellow Fever antibody positive<sup>3</sup> | 13 | 13 | 100% |
| Rheumatoid factors<sup>1</sup> | 8 | 8 | 100% |
| Hemolytic<sup>1</sup> | 11 | 11 | 100% |
| Icteric<sup>1</sup> | 11 | 11 | 100% |

<sup>1</sup> Tested at CT-Vacinas (UFMG).
<sup>2</sup> Tested at Laboratory of Respiratory Viruses and Measles (Fiocruz-RJ).
<sup>3</sup> Tested at Laboratory of Virology (USP).

### 3.4. Clinical performance evaluation of the rN-ELISA IgG kit

Forty-three individuals who tested positive for SARS-CoV-2 RNA through qRT-PCR were followed up for 30 days and assayed by the DPP rapid test and our rN-ELISA IgG kit. Of these, 19 presented a positive result for one or both serological test at the first blood collection and 24 patients were initially seronegative and then seroconverted during the follow up period. The median day of seroconversion for rN-ELISA IgG kit and DPP (IgG) was 14 days post PCR confirmation. All patients presented seroconversion as evaluated by the developed the rN-ELISA IgG kit within a maximum of 21 days after PCR confirmation (Supplementary materials, Table S4).

The positive agreement to PCR was evaluated using 157 serum samples, collected at different timepoints. Considering borderline results as negative, the rN-ELISA IgG kit displayed a 48% (32/66) positive agreement to PCR before 10 days post molecular confirmation, 82% (54/66) from 11 to 20 days, and 100.0% (25/25) after 21 days. The DPP rapid test (serological reference method) displayed a 47% (31/66) positive agreement to PCR before 10 days post confirmation, from 11 to 20 days post-PCR confirmation, the positive agreement was 73% (48/66) and after 21 days, the positive agreement to PCR was 84% (21/25) (Table 3).

**Table 3.**
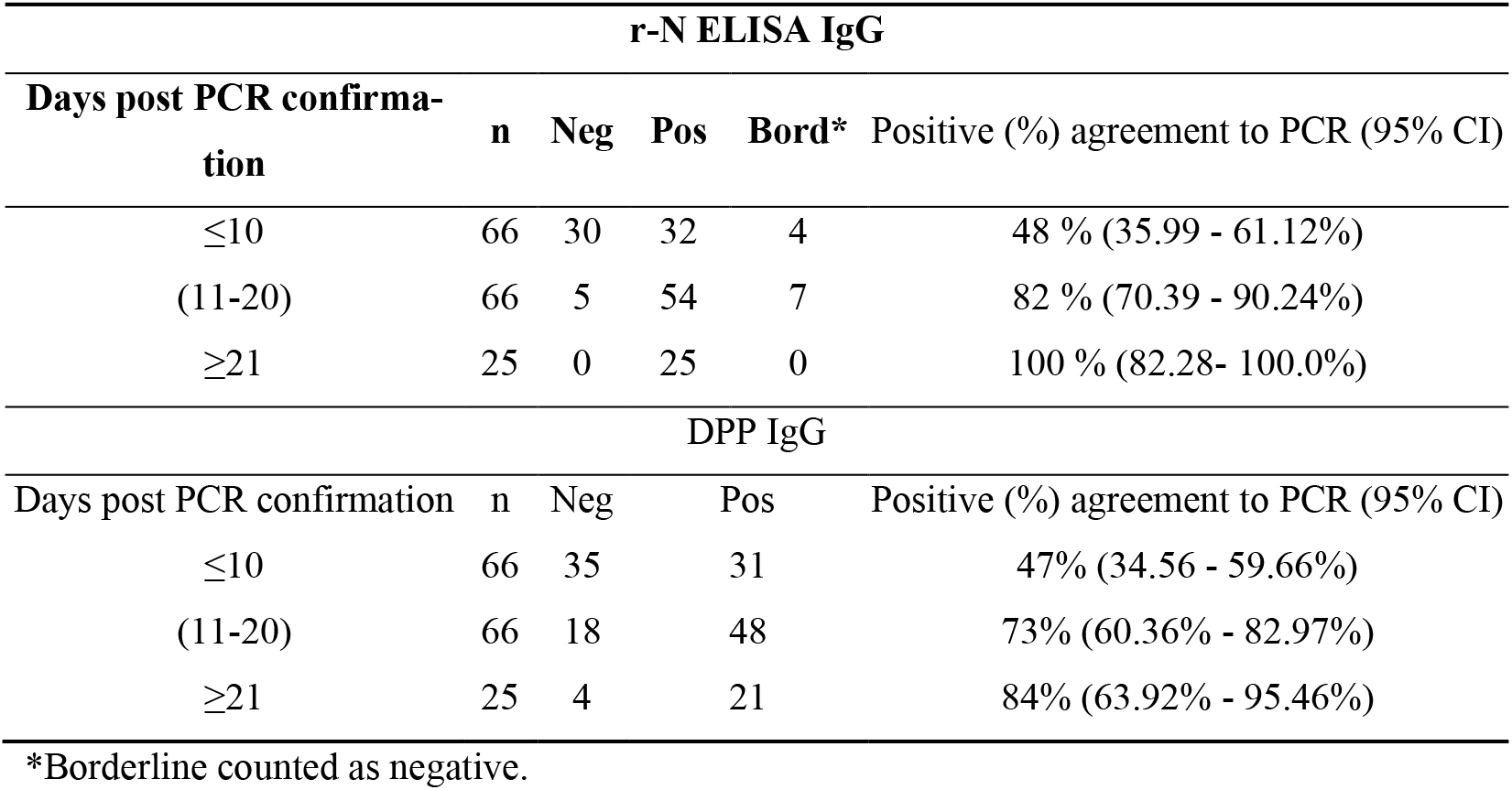
Positive agreement to PCR of anti-SARS-CoV-2 ELISA according to time.

A total of 261 samples were used to evaluate the agreement between the rN-ELISA IgG and DPP (serological reference), including samples at different times post-infection fom non-hospitalized individuals (n=157, from 43 patients after positive nasal swab PCR); hospitalized patients (n=19, from 11 individuals after positive nasal swab PCR); samples from healthy donors with a negative nasal swab PCR (n=23) and suspected cases (negative or no qPCR data, but that had contact with confirmed COVID-19 individuals, n=62) (Supplementary materials, Table S6 and S7). Considering samples with a positive result from both PCR and the serological reference test (DPP), the rN-ELISA IgG presented a sensitivity of 98.4% (CI:94.3-99.8%) (123/125, three borderline results, 0,8≤Index<1,1) and specificity of 100% (CI:88.8-100%) (31/32, one borderline result) based on PCR negative nasal swab or non-reagent DPP. Moreover, among the qPCR+/DPP-, the rN-ELISA IgG reacted against 20 samples. No false-positive tests (considering healthy donor with negative PCR) were observed. These results returned a K=0.775 (95% CI: 0.697-0.854), indicating a substantial agreement between both tests [18]. rN-ELISA IgG borderline results (n=19, three reactive by DPP and 16 non-reactive by DPP) were not included (Table 4).

**Table 4.**
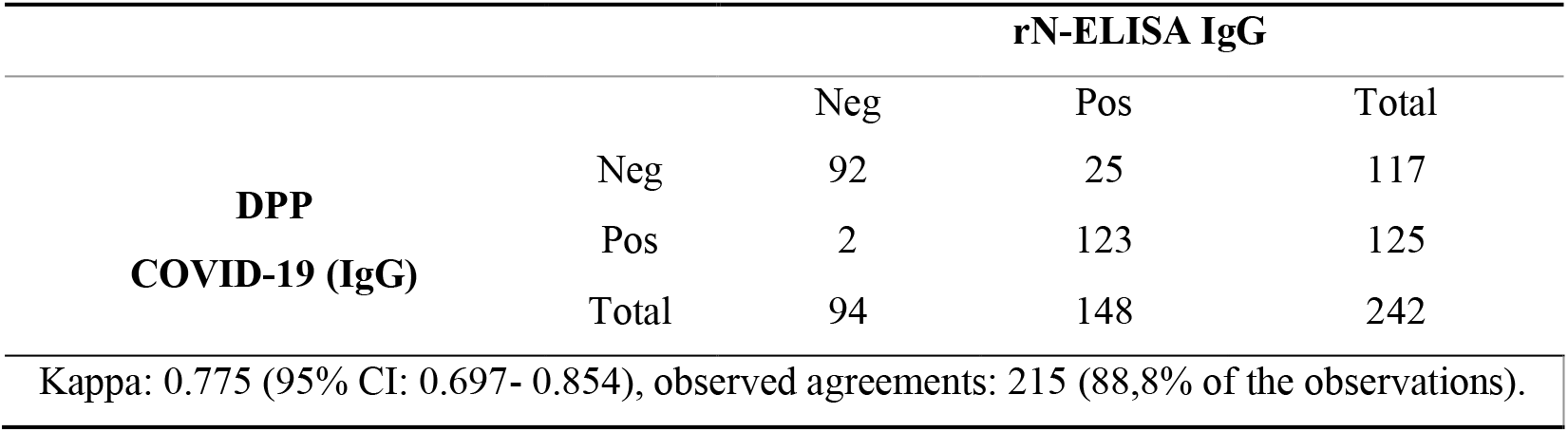
Agreement between the rN-ELISA and DPP (serological reference) to detect IgG against SARS-CoV-2.

### 3.6. Independent Clinical Agreement Validation Study

The rN-ELISA IgG kit was independently tested at three external laboratories: Laboratory of Virology, at the University of São Paulo (USP), Laboratory of Respiratory Viruses and Measles and Laboratory of Diagnostic Technology, both at the Oswaldo Cruz Foundation (Fiocruz-RJ). The test was validated against a panel of previously characterized samples consisting of SARS-CoV-2 PCR and/or antibody-positive samples (n= 168) and PCR negative (or prepandemic samples) (n=194). The rN-N ELISA sensitivity ranged from 84.6% to 95.7% and specificity from 97.7% to 100%. The test accuracy was higher than 90% in all three studies (Table 5).

**Table 5.** Independent Clinical Agreement Validation Study using rN-ELISA IgG.

|  | Positive samples | Negative samples | Sensitivity %<br>(CI 95%) | Specificity %<br>(CI 95%) | Accuracy %<br>(CI 95%) |
| --- | --- | --- | --- | --- | --- |
| Study<sup>1</sup> | qPCR + (n=52) | Before pandemic<br>(n=36) | 84.6 (72.5-92.0) % | 100(90.4-100.0)% | 93.2(85.8-97.5)% |
| Study<sup>2</sup> | qPCR + (n= 68) | PCR- (n=30) | 88.2 (78.5-93.9) % | 100(88.7-100.0)% | 91.3(82.8%-96.4)% |
| Study<sup>3</sup> | qPCR+/DPP+<br>(n=48) | Before pandemic<br>(n=128) | 95.7 (85.7-99.5) % | 97.7(93.3-99.5)% | 96.0(92.0-98.4)% |

<sup>1</sup>Laboratory of Virology (USP).
<sup>2</sup>Laboratory of Respiratory Viruses and Measles (Fiocruz-RJ).
<sup>3</sup>Laboratory of Diagnostic Technology (Fiocruz-RJ).

## 4. Discussion

Serological tests, such as Enzyme-linked immunosorbent assay (ELISA) and rapid tests available in Brazil are dependent on imported technology. This is also true for many developing countries where the pandemic has hit hard. The main imported raw material is the antigen, which makes the diagnosis more expensive, less accessible and subject to availability fluctuation due to international demands. Here, we described the development of a serological assay to detect IgG antibodies against SARS-CoV-2 infection. For the production of the recombinant antigen, a crucial step in terms of performance and costs, we used a prokaryotic system because, in terms of scale production, bacteria-made proteins present cost-effective advantages when compared to eukaryotic systems, including higher productivity and reduced costs [19]. Most commercial COVID-19 serological assays available to date use the Spike protein as the antigen [8,20–22]. However, to maintain its antigenic properties, this antigen must be produced in eukaryotic expression systems. The N protein has been described an immunodominant antigen generated during SASR-CoV-2 replication, and a superior antigenicity of N over S has been suggested [12,23,24]. Indeed, our rN-ELISA IgG kit based on the rN protein displayed similar performances when compared to tests from well-established manufacturers using the S1 antigen [25,26]. In fact, we also produced different versions of the S1 fragment of the S antigen to be used as antigens in ELISA, in *E. coli*; however, the recombinant S1 proteins presented lower yields, insolubility, and poorer performances to recognize SARS-CoV-2-positive sera when compared to the N protein (data not shown). Even when we compared our rN protein with a S1 commercial antigen, produced in a eucaryotic system (HEK293 cells), using the same ELISA components, the performance of the test using the N protein to recognize anti-SARS-CoV-2 IgGs was superior (data not shown). Furthermore, the developed rN-ELISA IgG kit presents improved sensitivity when compared to a commercially available test (DPP COVID-19 IgM/IgG, from Bio-Manguinhos, Fiocruz, Brazil).

Levels of antibodies in COVID-19 sera samples vary throughout the course of infection, and sensitivity data for serodiagnosis should be interpretated accordingly. In general, patients infected with SARS-CoV-2 display antibody responses between day 10 and 21 after the initial infection. Detection of antibodies in mild cases can take even longer (four weeks or more) and in a small number of cases, antibodies (i.e., IgM, IgG) were not detected at all (at least during the studies’ time scale). Based on the currently available data, the IgM and IgG antibodies to SARS-CoV-2 develop between 6–15 days post disease onset [6-9]. In our study, the median day of seroconversion for both EIE COVID-19 IgG kit and DPP (IgG) was 14 days post PCR confirmation.

Technology for the production of the rN-ELISA was transferred to the Laboratory of Diagnostic Technology, at Bio-Manguinhos Institute, where it was further developed into industrial batches. The rN-ELISA IgG is currently under the registration process at the Brazilian Sanitary and Healthcare Authority (ANVISA) and under field conditions external evaluation.

Considering the easy production of the antigen, the robustness of the test, the technical familiarity and the wide use of ELISA in clinical laboratory settings, and the poor accessibility in developing regions to more robust techniques such as PCR, this product represents an important addition to the currently available serodiagnosis toolbox for diagnosis of COVID-19.

## 5. Conclusions

Given that the developed rN-ELISA IgG kit presents high accuracy and robustness, it is amenable to be produced and can be considered as an excellent cost-benefit tool for detection of IgG antibodies in individual diagnosis or more broader applications, such as large-scale clinical studies and surveillance of SARS-CoV-2 infection.

## Data Availability

All data included in the manuscript is freely available.

## Supplementary Materials

**Table S1:** Demographic and clinical variables of mild symptomatic patients with confirmed SARS-CoV-2 infection from the internal validation (n=43). **Table S2:** Pre-pandemic samples used to calculate the cut-off during the rN-ELISA IgG internal validation. **Table S3:** Samples from healthy donors used to calculate the rN-ELISA IgG specificity during the internal validation. **Table S4:** Samples from qPCR positive patients used to calculate the rN-ELISA IgG sensitivity during the internal validation. **Table S5:** Patients from the seroconversion study tested by DPP and rN-ELISA IgG. **Table S6:** Negative and Suspected cases included to evaluate DPP and rN-ELISA agreement.

## Author Contributions

Conceptualization, F.F.B., R.T.G., S.M.R.T, A.P.S.M.F. and F.G.F.; methodology, F.F.B., R.T.G., S.M.R.T, L.A.F.A., N.C.S., A.P.S.M.F. and F.G.F.; validation, F.F.B., S.A.R.S., M.M.F., L.C.G., N.C.S., E.D.S., C.P.S., A.A, F. J.A., A.G.P.F, and E.L.D.; formal analysis, F.F.B., M.M.F., L.C.G., N.C.S., R.T.G., S.M.R.T, A.P.S.M.F. and F.G.F.; writing—original draft preparation, F.F.B., R.T.G., S.M.R.T, A.P.S.M.F. and F.G.F.; writing— review and editing, F.F.B., A.G.P.F, E.L.D., R.T.G., S.M.R.T, A.P.S.M.F. and F.G.F.; project administration, R.T.G., S.M.R.T, A.P.S.M.F. and F.G.F.; funding acquisition, R.T.G., S.M.R.T, A.P.S.M.F. and F.G.F. All authors have read and agreed to the published version of the manuscript.

## Acknowledgments

We thank all personnel at UFMG’s Vaccine Technology Center (CT-Vacinas) Belo Horizonte, Brazil, for their support and teamwork. We also thank Dr. Marilda A. M. T. de Siqueira, from Laboratory of Respiratory Viruses and Measles (Fiocruz-RJ, Brazil); teams from Laboratory of Virology (USP, Brazil); and Laboratory of Diagnostic Technology (Fiocruz-RJ, Brazil) for their support.

