## Supplemental data for "DEVELOPMENT AND VALIDATION OF AN ENZYME-LINKED IMMUNOASSAY KIT FOR DIAGNOSIS AND SURVEILLANCE OF COVID-19"

### Slide 1
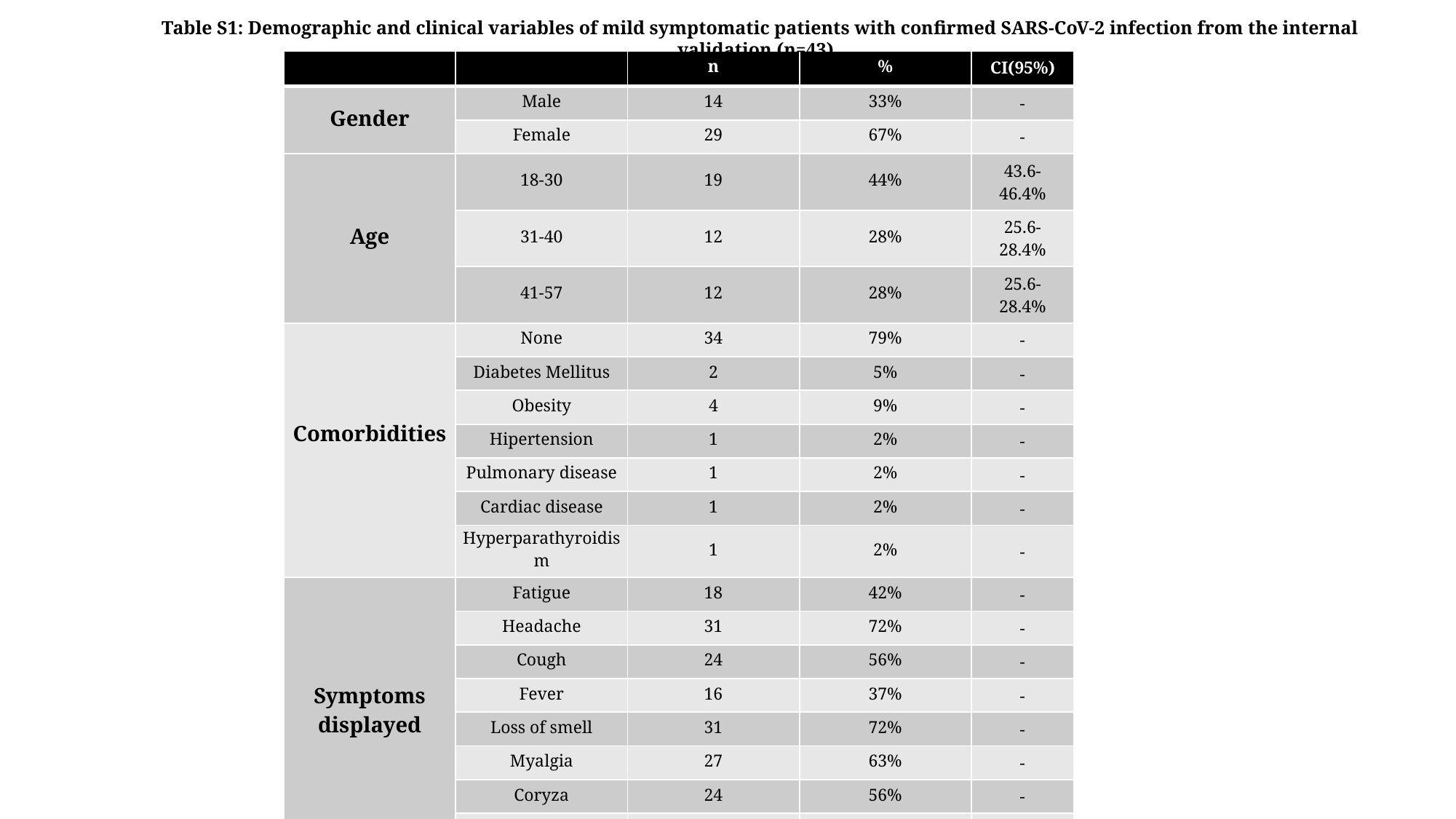

Table S1: Demographic and clinical variables of mild symptomatic patients with confirmed SARS-CoV-2 infection from the internal validation (n=43) .
| | | n | % | CI(95%) |
| --- | --- | --- | --- | --- |
| Gender | Male | 14 | 33% | - |
| | Female | 29 | 67% | - |
| Age | 18-30 | 19 | 44% | 43.6-46.4% |
| | 31-40 | 12 | 28% | 25.6-28.4% |
| | 41-57 | 12 | 28% | 25.6-28.4% |
| Comorbidities | None | 34 | 79% | - |
| | Diabetes Mellitus | 2 | 5% | - |
| | Obesity | 4 | 9% | - |
| | Hipertension | 1 | 2% | - |
| | Pulmonary disease | 1 | 2% | - |
| | Cardiac disease | 1 | 2% | - |
| | Hyperparathyroidism | 1 | 2% | - |
| Symptoms displayed | Fatigue | 18 | 42% | - |
| | Headache | 31 | 72% | - |
| | Cough | 24 | 56% | - |
| | Fever | 16 | 37% | - |
| | Loss of smell | 31 | 72% | - |
| | Myalgia | 27 | 63% | - |
| | Coryza | 24 | 56% | - |
| | Diarrhea | 17 | 40% | - |
| Period with symptoms | up to 5 days | 10 | 23% | 21.7-24.8% |
| | 6-10 days | 13 | 30% | 28.7-31.8% |
| | 11-16 days | 16 | 37% | 35.6-38.8% |
| | more than 20 days | 4 | 9% | 7.7-10.9% |

### Slide 2
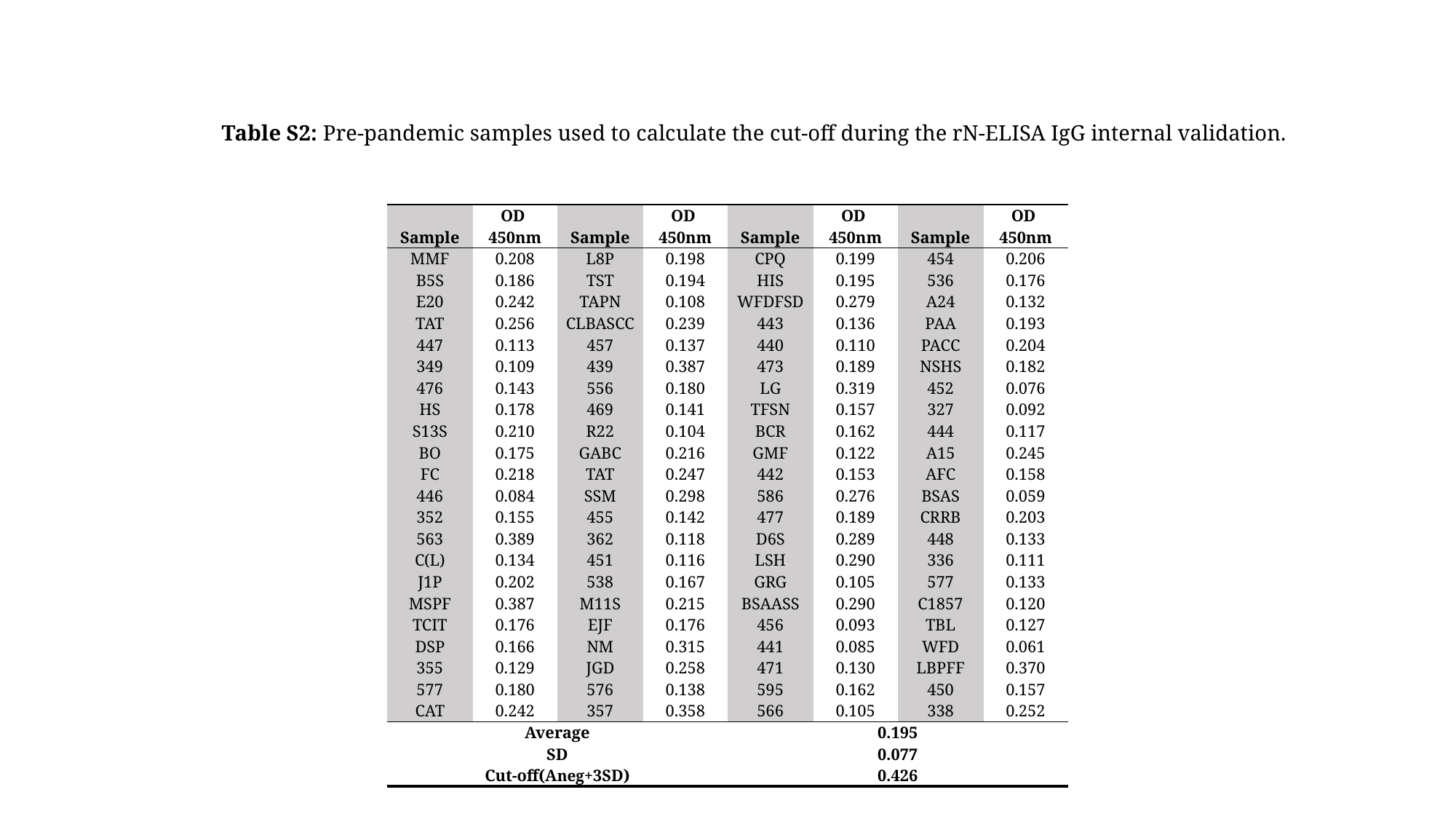

Table S2: Pre-pandemic samples used to calculate the cut-off during the rN-ELISA IgG internal validation.
| Sample | OD 450nm | Sample | OD 450nm | Sample | OD 450nm | Sample | OD 450nm |
| --- | --- | --- | --- | --- | --- | --- | --- |
| MMF | 0.208 | L8P | 0.198 | CPQ | 0.199 | 454 | 0.206 |
| B5S | 0.186 | TST | 0.194 | HIS | 0.195 | 536 | 0.176 |
| E20 | 0.242 | TAPN | 0.108 | WFDFSD | 0.279 | A24 | 0.132 |
| TAT | 0.256 | CLBASCC | 0.239 | 443 | 0.136 | PAA | 0.193 |
| 447 | 0.113 | 457 | 0.137 | 440 | 0.110 | PACC | 0.204 |
| 349 | 0.109 | 439 | 0.387 | 473 | 0.189 | NSHS | 0.182 |
| 476 | 0.143 | 556 | 0.180 | LG | 0.319 | 452 | 0.076 |
| HS | 0.178 | 469 | 0.141 | TFSN | 0.157 | 327 | 0.092 |
| S13S | 0.210 | R22 | 0.104 | BCR | 0.162 | 444 | 0.117 |
| BO | 0.175 | GABC | 0.216 | GMF | 0.122 | A15 | 0.245 |
| FC | 0.218 | TAT | 0.247 | 442 | 0.153 | AFC | 0.158 |
| 446 | 0.084 | SSM | 0.298 | 586 | 0.276 | BSAS | 0.059 |
| 352 | 0.155 | 455 | 0.142 | 477 | 0.189 | CRRB | 0.203 |
| 563 | 0.389 | 362 | 0.118 | D6S | 0.289 | 448 | 0.133 |
| C(L) | 0.134 | 451 | 0.116 | LSH | 0.290 | 336 | 0.111 |
| J1P | 0.202 | 538 | 0.167 | GRG | 0.105 | 577 | 0.133 |
| MSPF | 0.387 | M11S | 0.215 | BSAASS | 0.290 | C1857 | 0.120 |
| TCIT | 0.176 | EJF | 0.176 | 456 | 0.093 | TBL | 0.127 |
| DSP | 0.166 | NM | 0.315 | 441 | 0.085 | WFD | 0.061 |
| 355 | 0.129 | JGD | 0.258 | 471 | 0.130 | LBPFF | 0.370 |
| 577 | 0.180 | 576 | 0.138 | 595 | 0.162 | 450 | 0.157 |
| CAT | 0.242 | 357 | 0.358 | 566 | 0.105 | 338 | 0.252 |
| Average | | | | 0.195 | | | |
| SD | | | | 0.077 | | | |
| Cut-off(Aneg+3SD) | | | | 0.426 | | | |

### Slide 3
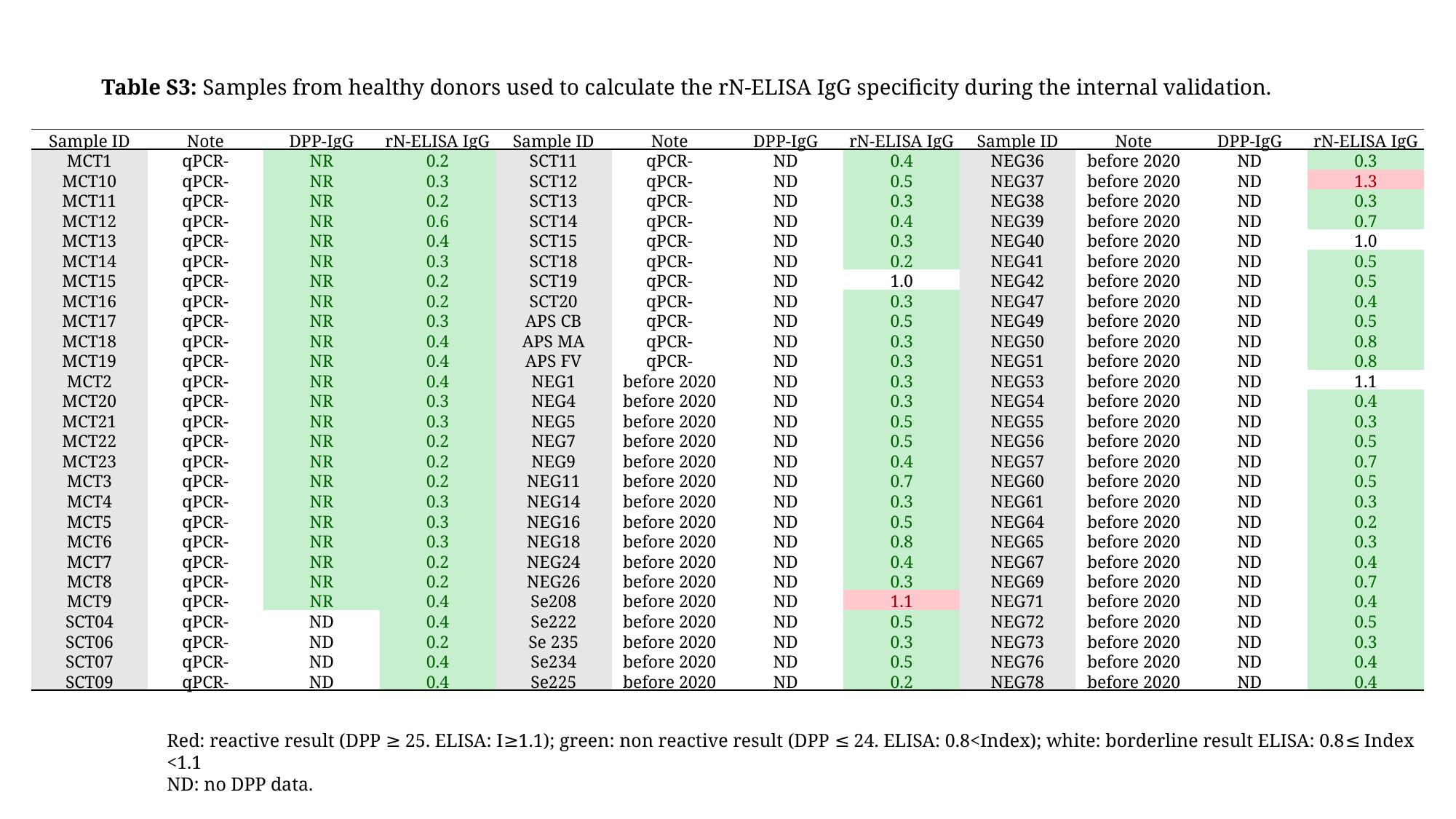

Table S3: Samples from healthy donors used to calculate the rN-ELISA IgG specificity during the internal validation.
| Sample ID | Note | DPP-IgG | rN-ELISA IgG | Sample ID | Note | DPP-IgG | rN-ELISA IgG | Sample ID | Note | DPP-IgG | rN-ELISA IgG |
| --- | --- | --- | --- | --- | --- | --- | --- | --- | --- | --- | --- |
| MCT1 | qPCR- | NR | 0.2 | SCT11 | qPCR- | ND | 0.4 | NEG36 | before 2020 | ND | 0.3 |
| MCT10 | qPCR- | NR | 0.3 | SCT12 | qPCR- | ND | 0.5 | NEG37 | before 2020 | ND | 1.3 |
| MCT11 | qPCR- | NR | 0.2 | SCT13 | qPCR- | ND | 0.3 | NEG38 | before 2020 | ND | 0.3 |
| MCT12 | qPCR- | NR | 0.6 | SCT14 | qPCR- | ND | 0.4 | NEG39 | before 2020 | ND | 0.7 |
| MCT13 | qPCR- | NR | 0.4 | SCT15 | qPCR- | ND | 0.3 | NEG40 | before 2020 | ND | 1.0 |
| MCT14 | qPCR- | NR | 0.3 | SCT18 | qPCR- | ND | 0.2 | NEG41 | before 2020 | ND | 0.5 |
| MCT15 | qPCR- | NR | 0.2 | SCT19 | qPCR- | ND | 1.0 | NEG42 | before 2020 | ND | 0.5 |
| MCT16 | qPCR- | NR | 0.2 | SCT20 | qPCR- | ND | 0.3 | NEG47 | before 2020 | ND | 0.4 |
| MCT17 | qPCR- | NR | 0.3 | APS CB | qPCR- | ND | 0.5 | NEG49 | before 2020 | ND | 0.5 |
| MCT18 | qPCR- | NR | 0.4 | APS MA | qPCR- | ND | 0.3 | NEG50 | before 2020 | ND | 0.8 |
| MCT19 | qPCR- | NR | 0.4 | APS FV | qPCR- | ND | 0.3 | NEG51 | before 2020 | ND | 0.8 |
| MCT2 | qPCR- | NR | 0.4 | NEG1 | before 2020 | ND | 0.3 | NEG53 | before 2020 | ND | 1.1 |
| MCT20 | qPCR- | NR | 0.3 | NEG4 | before 2020 | ND | 0.3 | NEG54 | before 2020 | ND | 0.4 |
| MCT21 | qPCR- | NR | 0.3 | NEG5 | before 2020 | ND | 0.5 | NEG55 | before 2020 | ND | 0.3 |
| MCT22 | qPCR- | NR | 0.2 | NEG7 | before 2020 | ND | 0.5 | NEG56 | before 2020 | ND | 0.5 |
| MCT23 | qPCR- | NR | 0.2 | NEG9 | before 2020 | ND | 0.4 | NEG57 | before 2020 | ND | 0.7 |
| MCT3 | qPCR- | NR | 0.2 | NEG11 | before 2020 | ND | 0.7 | NEG60 | before 2020 | ND | 0.5 |
| MCT4 | qPCR- | NR | 0.3 | NEG14 | before 2020 | ND | 0.3 | NEG61 | before 2020 | ND | 0.3 |
| MCT5 | qPCR- | NR | 0.3 | NEG16 | before 2020 | ND | 0.5 | NEG64 | before 2020 | ND | 0.2 |
| MCT6 | qPCR- | NR | 0.3 | NEG18 | before 2020 | ND | 0.8 | NEG65 | before 2020 | ND | 0.3 |
| MCT7 | qPCR- | NR | 0.2 | NEG24 | before 2020 | ND | 0.4 | NEG67 | before 2020 | ND | 0.4 |
| MCT8 | qPCR- | NR | 0.2 | NEG26 | before 2020 | ND | 0.3 | NEG69 | before 2020 | ND | 0.7 |
| MCT9 | qPCR- | NR | 0.4 | Se208 | before 2020 | ND | 1.1 | NEG71 | before 2020 | ND | 0.4 |
| SCT04 | qPCR- | ND | 0.4 | Se222 | before 2020 | ND | 0.5 | NEG72 | before 2020 | ND | 0.5 |
| SCT06 | qPCR- | ND | 0.2 | Se 235 | before 2020 | ND | 0.3 | NEG73 | before 2020 | ND | 0.3 |
| SCT07 | qPCR- | ND | 0.4 | Se234 | before 2020 | ND | 0.5 | NEG76 | before 2020 | ND | 0.4 |
| SCT09 | qPCR- | ND | 0.4 | Se225 | before 2020 | ND | 0.2 | NEG78 | before 2020 | ND | 0.4 |
Red: reactive result (DPP ≥ 25. ELISA: I≥1.1); green: non reactive result (DPP ≤ 24. ELISA: 0.8<Index); white: borderline result ELISA: 0.8≤ Index <1.1
ND: no DPP data.

### Slide 4
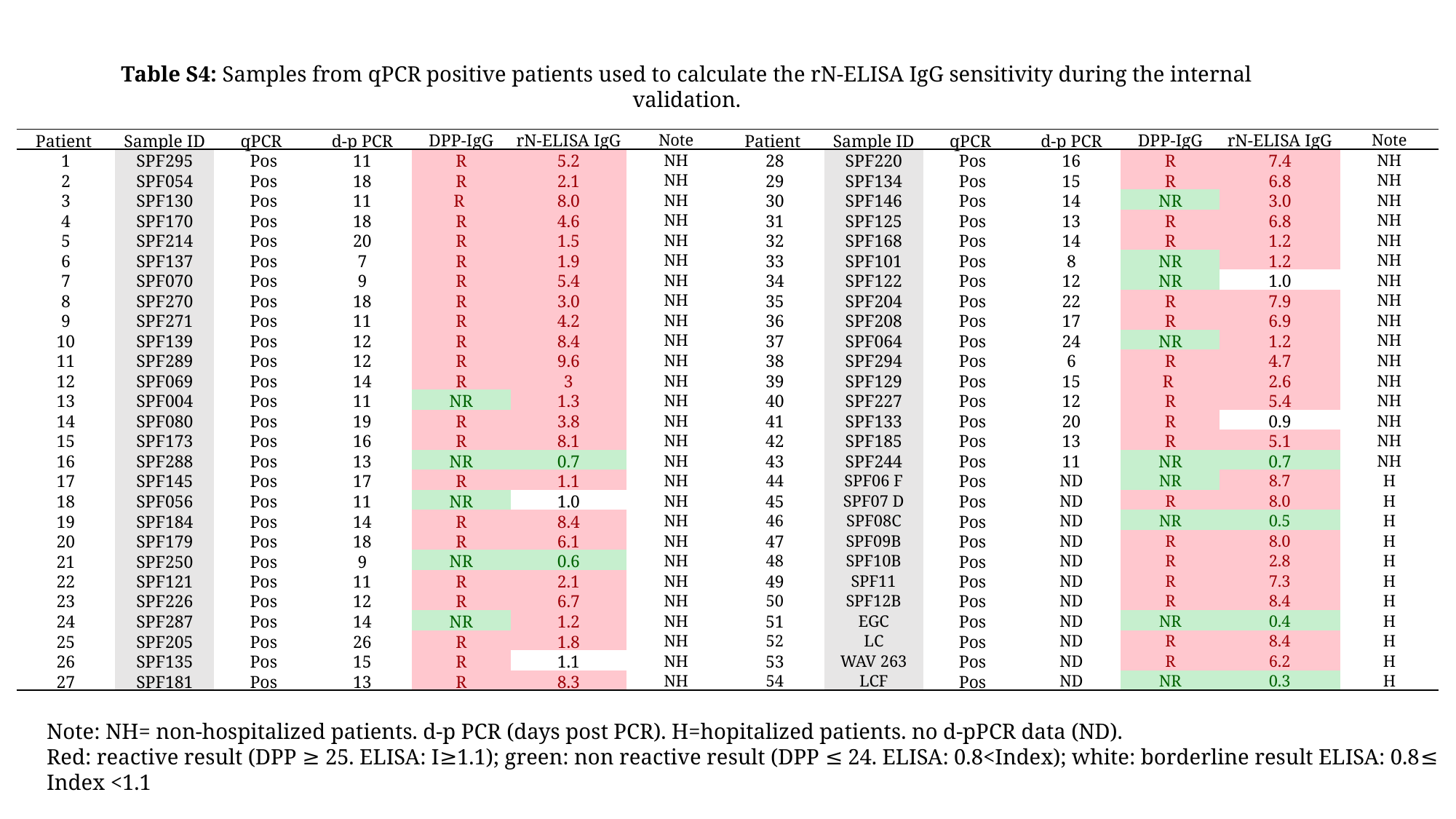

Table S4: Samples from qPCR positive patients used to calculate the rN-ELISA IgG sensitivity during the internal validation.
| Patient | Sample ID | qPCR | d-p PCR | DPP-IgG | rN-ELISA IgG | Note | Patient | Sample ID | qPCR | d-p PCR | DPP-IgG | rN-ELISA IgG | Note |
| --- | --- | --- | --- | --- | --- | --- | --- | --- | --- | --- | --- | --- | --- |
| 1 | SPF295 | Pos | 11 | R | 5.2 | NH | 28 | SPF220 | Pos | 16 | R | 7.4 | NH |
| 2 | SPF054 | Pos | 18 | R | 2.1 | NH | 29 | SPF134 | Pos | 15 | R | 6.8 | NH |
| 3 | SPF130 | Pos | 11 | R | 8.0 | NH | 30 | SPF146 | Pos | 14 | NR | 3.0 | NH |
| 4 | SPF170 | Pos | 18 | R | 4.6 | NH | 31 | SPF125 | Pos | 13 | R | 6.8 | NH |
| 5 | SPF214 | Pos | 20 | R | 1.5 | NH | 32 | SPF168 | Pos | 14 | R | 1.2 | NH |
| 6 | SPF137 | Pos | 7 | R | 1.9 | NH | 33 | SPF101 | Pos | 8 | NR | 1.2 | NH |
| 7 | SPF070 | Pos | 9 | R | 5.4 | NH | 34 | SPF122 | Pos | 12 | NR | 1.0 | NH |
| 8 | SPF270 | Pos | 18 | R | 3.0 | NH | 35 | SPF204 | Pos | 22 | R | 7.9 | NH |
| 9 | SPF271 | Pos | 11 | R | 4.2 | NH | 36 | SPF208 | Pos | 17 | R | 6.9 | NH |
| 10 | SPF139 | Pos | 12 | R | 8.4 | NH | 37 | SPF064 | Pos | 24 | NR | 1.2 | NH |
| 11 | SPF289 | Pos | 12 | R | 9.6 | NH | 38 | SPF294 | Pos | 6 | R | 4.7 | NH |
| 12 | SPF069 | Pos | 14 | R | 3 | NH | 39 | SPF129 | Pos | 15 | R | 2.6 | NH |
| 13 | SPF004 | Pos | 11 | NR | 1.3 | NH | 40 | SPF227 | Pos | 12 | R | 5.4 | NH |
| 14 | SPF080 | Pos | 19 | R | 3.8 | NH | 41 | SPF133 | Pos | 20 | R | 0.9 | NH |
| 15 | SPF173 | Pos | 16 | R | 8.1 | NH | 42 | SPF185 | Pos | 13 | R | 5.1 | NH |
| 16 | SPF288 | Pos | 13 | NR | 0.7 | NH | 43 | SPF244 | Pos | 11 | NR | 0.7 | NH |
| 17 | SPF145 | Pos | 17 | R | 1.1 | NH | 44 | SPF06 F | Pos | ND | NR | 8.7 | H |
| 18 | SPF056 | Pos | 11 | NR | 1.0 | NH | 45 | SPF07 D | Pos | ND | R | 8.0 | H |
| 19 | SPF184 | Pos | 14 | R | 8.4 | NH | 46 | SPF08C | Pos | ND | NR | 0.5 | H |
| 20 | SPF179 | Pos | 18 | R | 6.1 | NH | 47 | SPF09B | Pos | ND | R | 8.0 | H |
| 21 | SPF250 | Pos | 9 | NR | 0.6 | NH | 48 | SPF10B | Pos | ND | R | 2.8 | H |
| 22 | SPF121 | Pos | 11 | R | 2.1 | NH | 49 | SPF11 | Pos | ND | R | 7.3 | H |
| 23 | SPF226 | Pos | 12 | R | 6.7 | NH | 50 | SPF12B | Pos | ND | R | 8.4 | H |
| 24 | SPF287 | Pos | 14 | NR | 1.2 | NH | 51 | EGC | Pos | ND | NR | 0.4 | H |
| 25 | SPF205 | Pos | 26 | R | 1.8 | NH | 52 | LC | Pos | ND | R | 8.4 | H |
| 26 | SPF135 | Pos | 15 | R | 1.1 | NH | 53 | WAV 263 | Pos | ND | R | 6.2 | H |
| 27 | SPF181 | Pos | 13 | R | 8.3 | NH | 54 | LCF | Pos | ND | NR | 0.3 | H |
Note: NH= non-hospitalized patients. d-p PCR (days post PCR). H=hopitalized patients. no d-pPCR data (ND).
Red: reactive result (DPP ≥ 25. ELISA: I≥1.1); green: non reactive result (DPP ≤ 24. ELISA: 0.8<Index); white: borderline result ELISA: 0.8≤ Index <1.1

### Slide 5
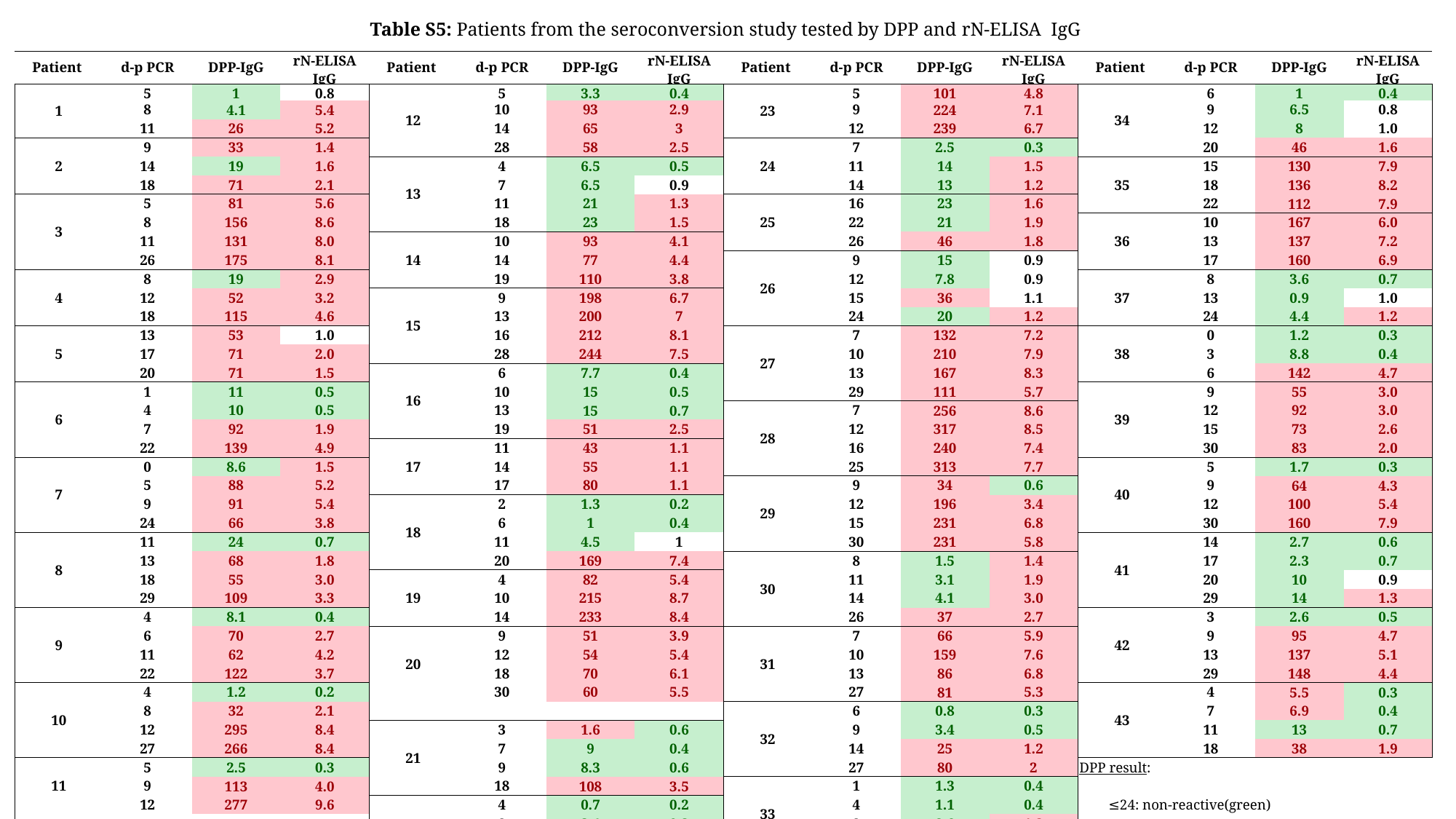

Table S5: Patients from the seroconversion study tested by DPP and rN-ELISA IgG
| Patient | d-p PCR | DPP-IgG | rN-ELISA IgG | Patient | d-p PCR | DPP-IgG | rN-ELISA IgG | Patient | d-p PCR | DPP-IgG | rN-ELISA IgG | Patient | d-p PCR | DPP-IgG | rN-ELISA IgG |
| --- | --- | --- | --- | --- | --- | --- | --- | --- | --- | --- | --- | --- | --- | --- | --- |
| 1 | 5 | 1 | 0.8 | 12 | 5 | 3.3 | 0.4 | 23 | 5 | 101 | 4.8 | 34 | 6 | 1 | 0.4 |
| | 8 | 4.1 | 5.4 | | 10 | 93 | 2.9 | | 9 | 224 | 7.1 | | 9 | 6.5 | 0.8 |
| | 11 | 26 | 5.2 | | 14 | 65 | 3 | | 12 | 239 | 6.7 | | 12 | 8 | 1.0 |
| 2 | 9 | 33 | 1.4 | | 28 | 58 | 2.5 | 24 | 7 | 2.5 | 0.3 | | 20 | 46 | 1.6 |
| | 14 | 19 | 1.6 | 13 | 4 | 6.5 | 0.5 | | 11 | 14 | 1.5 | 35 | 15 | 130 | 7.9 |
| | 18 | 71 | 2.1 | | 7 | 6.5 | 0.9 | | 14 | 13 | 1.2 | | 18 | 136 | 8.2 |
| 3 | 5 | 81 | 5.6 | | 11 | 21 | 1.3 | 25 | 16 | 23 | 1.6 | | 22 | 112 | 7.9 |
| | 8 | 156 | 8.6 | | 18 | 23 | 1.5 | | 22 | 21 | 1.9 | 36 | 10 | 167 | 6.0 |
| | 11 | 131 | 8.0 | 14 | 10 | 93 | 4.1 | | 26 | 46 | 1.8 | | 13 | 137 | 7.2 |
| | 26 | 175 | 8.1 | | 14 | 77 | 4.4 | 26 | 9 | 15 | 0.9 | | 17 | 160 | 6.9 |
| 4 | 8 | 19 | 2.9 | | 19 | 110 | 3.8 | | 12 | 7.8 | 0.9 | 37 | 8 | 3.6 | 0.7 |
| | 12 | 52 | 3.2 | 15 | 9 | 198 | 6.7 | | 15 | 36 | 1.1 | | 13 | 0.9 | 1.0 |
| | 18 | 115 | 4.6 | | 13 | 200 | 7 | | 24 | 20 | 1.2 | | 24 | 4.4 | 1.2 |
| 5 | 13 | 53 | 1.0 | | 16 | 212 | 8.1 | 27 | 7 | 132 | 7.2 | 38 | 0 | 1.2 | 0.3 |
| | 17 | 71 | 2.0 | | 28 | 244 | 7.5 | | 10 | 210 | 7.9 | | 3 | 8.8 | 0.4 |
| | 20 | 71 | 1.5 | 16 | 6 | 7.7 | 0.4 | | 13 | 167 | 8.3 | | 6 | 142 | 4.7 |
| 6 | 1 | 11 | 0.5 | | 10 | 15 | 0.5 | | 29 | 111 | 5.7 | 39 | 9 | 55 | 3.0 |
| | 4 | 10 | 0.5 | | 13 | 15 | 0.7 | 28 | 7 | 256 | 8.6 | | 12 | 92 | 3.0 |
| | 7 | 92 | 1.9 | | 19 | 51 | 2.5 | | 12 | 317 | 8.5 | | 15 | 73 | 2.6 |
| | 22 | 139 | 4.9 | 17 | 11 | 43 | 1.1 | | 16 | 240 | 7.4 | | 30 | 83 | 2.0 |
| 7 | 0 | 8.6 | 1.5 | | 14 | 55 | 1.1 | | 25 | 313 | 7.7 | 40 | 5 | 1.7 | 0.3 |
| | 5 | 88 | 5.2 | | 17 | 80 | 1.1 | 29 | 9 | 34 | 0.6 | | 9 | 64 | 4.3 |
| | 9 | 91 | 5.4 | 18 | 2 | 1.3 | 0.2 | | 12 | 196 | 3.4 | | 12 | 100 | 5.4 |
| | 24 | 66 | 3.8 | | 6 | 1 | 0.4 | | 15 | 231 | 6.8 | | 30 | 160 | 7.9 |
| 8 | 11 | 24 | 0.7 | | 11 | 4.5 | 1 | | 30 | 231 | 5.8 | 41 | 14 | 2.7 | 0.6 |
| | 13 | 68 | 1.8 | | 20 | 169 | 7.4 | 30 | 8 | 1.5 | 1.4 | | 17 | 2.3 | 0.7 |
| | 18 | 55 | 3.0 | 19 | 4 | 82 | 5.4 | | 11 | 3.1 | 1.9 | | 20 | 10 | 0.9 |
| | 29 | 109 | 3.3 | | 10 | 215 | 8.7 | | 14 | 4.1 | 3.0 | | 29 | 14 | 1.3 |
| 9 | 4 | 8.1 | 0.4 | | 14 | 233 | 8.4 | | 26 | 37 | 2.7 | 42 | 3 | 2.6 | 0.5 |
| | 6 | 70 | 2.7 | 20 | 9 | 51 | 3.9 | 31 | 7 | 66 | 5.9 | | 9 | 95 | 4.7 |
| | 11 | 62 | 4.2 | | 12 | 54 | 5.4 | | 10 | 159 | 7.6 | | 13 | 137 | 5.1 |
| | 22 | 122 | 3.7 | | 18 | 70 | 6.1 | | 13 | 86 | 6.8 | | 29 | 148 | 4.4 |
| 10 | 4 | 1.2 | 0.2 | | 30 | 60 | 5.5 | | 27 | 81 | 5.3 | 43 | 4 | 5.5 | 0.3 |
| | 8 | 32 | 2.1 | | | | | 32 | 6 | 0.8 | 0.3 | | 7 | 6.9 | 0.4 |
| | 12 | 295 | 8.4 | 21 | 3 | 1.6 | 0.6 | | 9 | 3.4 | 0.5 | | 11 | 13 | 0.7 |
| | 27 | 266 | 8.4 | | 7 | 9 | 0.4 | | 14 | 25 | 1.2 | | 18 | 38 | 1.9 |
| 11 | 5 | 2.5 | 0.3 | | 9 | 8.3 | 0.6 | | 27 | 80 | 2 | DPP result: ≤24: non-reactive(green) ≥25: reactive ELISA result: 0.8<I: non reactive (green) 0.8≤ I <1.1: borderline(white) I≥1.1: reactive (red) | | | |
| | 9 | 113 | 4.0 | | 18 | 108 | 3.5 | 33 | 1 | 1.3 | 0.4 | | | | |
| | 12 | 277 | 9.6 | 22 | 4 | 0.7 | 0.2 | | 4 | 1.1 | 0.4 | | | | |
| | | | | | 8 | 2.4 | 0.3 | | 8 | 8.6 | 1.2 | | | | |
| | | | | | 11 | 88 | 2.1 | | 15 | 146 | 5.9 | | | | |
| | | | | | 27 | 279 | 8.3 | | | | | | | | |

### Slide 6
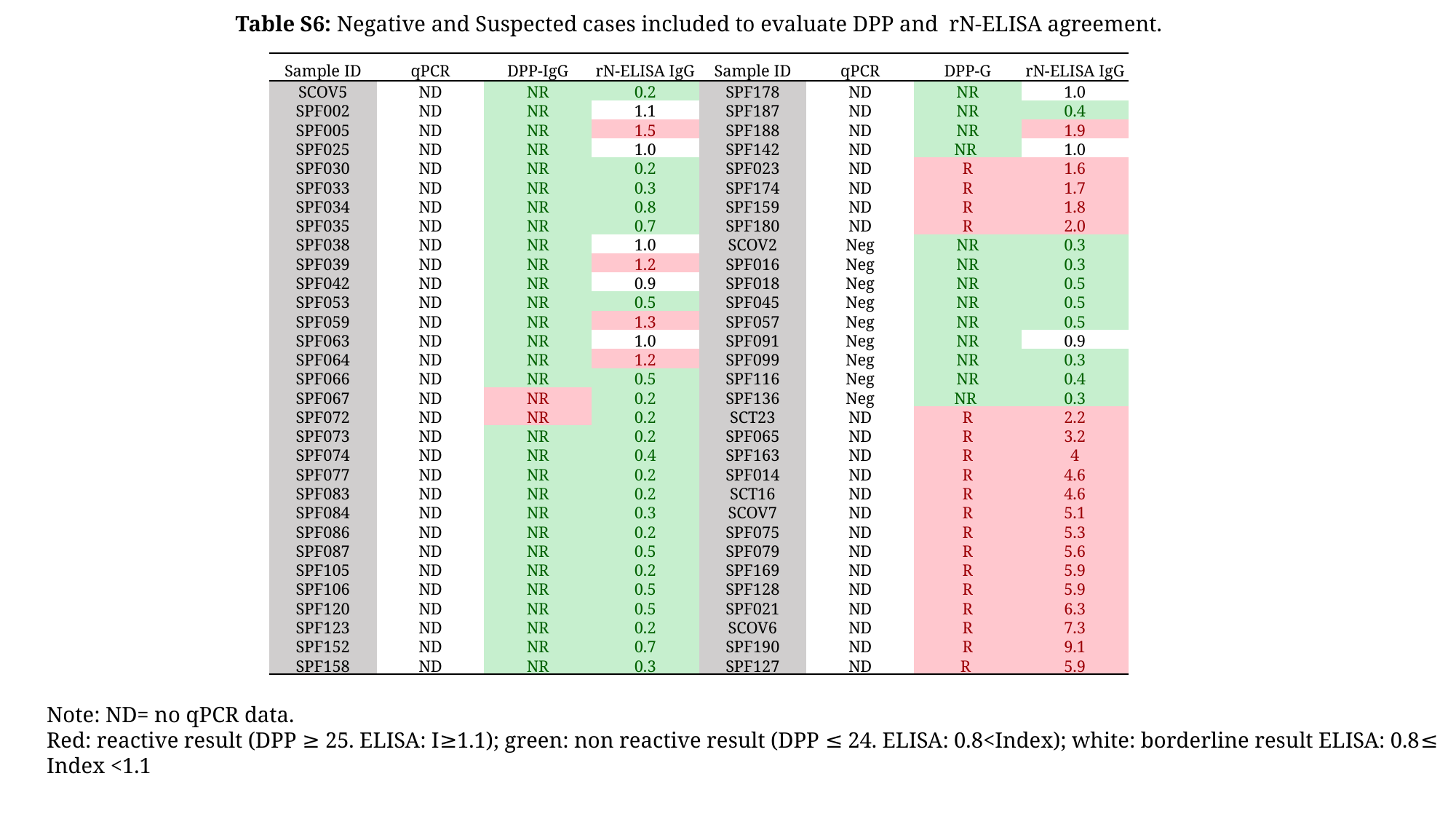

Table S6: Negative and Suspected cases included to evaluate DPP and  rN-ELISA agreement.
| Sample ID | qPCR | DPP-IgG | rN-ELISA IgG | Sample ID | qPCR | DPP-G | rN-ELISA IgG |
| --- | --- | --- | --- | --- | --- | --- | --- |
| SCOV5 | ND | NR | 0.2 | SPF178 | ND | NR | 1.0 |
| SPF002 | ND | NR | 1.1 | SPF187 | ND | NR | 0.4 |
| SPF005 | ND | NR | 1.5 | SPF188 | ND | NR | 1.9 |
| SPF025 | ND | NR | 1.0 | SPF142 | ND | NR | 1.0 |
| SPF030 | ND | NR | 0.2 | SPF023 | ND | R | 1.6 |
| SPF033 | ND | NR | 0.3 | SPF174 | ND | R | 1.7 |
| SPF034 | ND | NR | 0.8 | SPF159 | ND | R | 1.8 |
| SPF035 | ND | NR | 0.7 | SPF180 | ND | R | 2.0 |
| SPF038 | ND | NR | 1.0 | SCOV2 | Neg | NR | 0.3 |
| SPF039 | ND | NR | 1.2 | SPF016 | Neg | NR | 0.3 |
| SPF042 | ND | NR | 0.9 | SPF018 | Neg | NR | 0.5 |
| SPF053 | ND | NR | 0.5 | SPF045 | Neg | NR | 0.5 |
| SPF059 | ND | NR | 1.3 | SPF057 | Neg | NR | 0.5 |
| SPF063 | ND | NR | 1.0 | SPF091 | Neg | NR | 0.9 |
| SPF064 | ND | NR | 1.2 | SPF099 | Neg | NR | 0.3 |
| SPF066 | ND | NR | 0.5 | SPF116 | Neg | NR | 0.4 |
| SPF067 | ND | NR | 0.2 | SPF136 | Neg | NR | 0.3 |
| SPF072 | ND | NR | 0.2 | SCT23 | ND | R | 2.2 |
| SPF073 | ND | NR | 0.2 | SPF065 | ND | R | 3.2 |
| SPF074 | ND | NR | 0.4 | SPF163 | ND | R | 4 |
| SPF077 | ND | NR | 0.2 | SPF014 | ND | R | 4.6 |
| SPF083 | ND | NR | 0.2 | SCT16 | ND | R | 4.6 |
| SPF084 | ND | NR | 0.3 | SCOV7 | ND | R | 5.1 |
| SPF086 | ND | NR | 0.2 | SPF075 | ND | R | 5.3 |
| SPF087 | ND | NR | 0.5 | SPF079 | ND | R | 5.6 |
| SPF105 | ND | NR | 0.2 | SPF169 | ND | R | 5.9 |
| SPF106 | ND | NR | 0.5 | SPF128 | ND | R | 5.9 |
| SPF120 | ND | NR | 0.5 | SPF021 | ND | R | 6.3 |
| SPF123 | ND | NR | 0.2 | SCOV6 | ND | R | 7.3 |
| SPF152 | ND | NR | 0.7 | SPF190 | ND | R | 9.1 |
| SPF158 | ND | NR | 0.3 | SPF127 | ND | R | 5.9 |
Note: ND= no qPCR data.
Red: reactive result (DPP ≥ 25. ELISA: I≥1.1); green: non reactive result (DPP ≤ 24. ELISA: 0.8<Index); white: borderline result ELISA: 0.8≤ Index <1.1
